# Incidence and Predictors of Recurrent Pericarditis in Systemic Lupus Erythematosus

**DOI:** 10.1101/2024.06.09.24308628

**Authors:** Yoo Jin Kim, Jana Lovell, Alaa Diab, Laurence S. Magder, Daniel Goldman, Michelle Petri, Andrea Fava, Luigi Adamo

## Abstract

**IMPORTANCE:** Pericarditis is the most common cardiac manifestation of Systemic Lupus Erythematosus (SLE) and known to recur among patients. Yet, the prevalence and risk factors of recurrent pericarditis in SLE patients are unknown.

**OBJECTIVE:** Determine the frequency and risk factors for the recurrence of pericarditis in patients with SLE.

**DESIGN:** Retrospective analysis of a well-characterized, prospective cohort of SLE patients enrolled between 1988 and 2023.

**SETTING:** A single-center cohort study of a diverse group of SLE patients treated at a tertiary medical center.

**PARTICIPANTS:** Patients diagnosed with pericarditis (n=590) among those enrolled in the Hopkins Lupus Cohort (n=2931).

**MAIN OUTCOME:** Re-occurrence of pericarditis. The SELENA revision of the SLE Disease Activity Index (SLEDAI) was used to define pericarditis. Clinical information was examined for all follow-up encounters after the first episode of pericarditis. Pericarditis that occurred at least six weeks after the first recorded episode was defined as “Recurrent”.

**RESULTS:** Of 2931 patients within the cohort, 590 had a history of pericarditis. In 3.4% of patients, the diagnosis of pericarditis was confirmed via electrocardiogram (EKG) or dedicated imaging, with 100% concordance between clinical and data-based diagnoses. During a median follow-up of 7 years (IQR: 3 – 14), 20% (n=120) of patients experienced recurrent pericarditis (recurrence rate ≈ 0.05 per person-year of follow-up). Most patients (51%) experienced only one recurrence, whereas 49% had ≥2 recurrences. In multivariate analysis, predictors of recurrence included younger age (≥60 years vs. <40, RR 0.11 (0.04, 0.32), *P* <.001), treatment with prednisone (≥20 mg vs. 0, RR 1.99 (1.17, 3.40), *P* = 0.012), active SLE disease (SLEDAI ≥3 vs. 0, RR 1.55 (1.21, 2.00), *P* <.001), and time since initial episode (3-10 years vs. <1, RR 0.32 (0.20, 0.52), *P* <.001).

**CONCLUSION AND RELEVANCE:** Recurrence is more likely to occur within one year of the onset of pericarditis, and younger patients and those with uncontrolled disease are at greater risk of recurrence. As in the general population, oral prednisone therapy is associated with a higher chance of recurrence in SLE patients, with a dose-dependent effect. These findings set the basis for future studies to define optimal treatment for recurrent pericarditis in SLE patients and suggest that oral corticosteroids should be avoided when treating pericarditis.

## INTRODUCTION

Pericarditis, defined as inflammation of the serosal sac that surrounds the myocardium, is the most common cardiac manifestation of systemic lupus erythematosus (SLE) and noted in about 20% of patients.^1–3^ Patients can experience a spectrum of symptoms, ranging from mild chest pain exacerbated by lying flat that is improved by leaning forward, to debilitating symptoms of severe chest pain and dyspnea. Importantly, pericarditis is associated with complications that include recurrence, myocarditis, and pericardial effusion.

In the general population, pericarditis recurs in approximately 30% of cases.^4^ Despite the significant rate of recurrence, the mechanisms predisposing a subset of individuals to recurrence remains unclear. Caforio and colleagues demonstrated the presence of serum anti-heart and anti-intercalated-disk autoantibodies in patients with recurrent pericarditis.^5^ Furthermore, multiple clinical trials have demonstrated the efficacy of immunosuppressive agents, such as corticosteroids, in treating recurrent pericarditis, suggesting an immune-mediated mechanism.^5–8^ Recent pivotal studies further support this hypothesis, particularly implicating interleukin-1, a pro-inflammatory cytokine of the innate immune system, in the pathophysiology of recurrent pericarditis. Inhibitors targeting this pathway have proven effective in treating idiopathic recurrent pericarditis across multiple randomized clinical trials.^7–14^

Given the broad immune dysregulation associated with SLE, patients may face greater risk for recurrence due to possible overlapping immune-mediated mechanisms. However, the rate of recurrence of pericarditis in SLE patients, its risk factors, and its association with various treatments have not been characterized. To address this knowledge gap, we analyzed the Hopkins Lupus Cohort, a large and well-characterized longitudinal cohort of SLE patients.

## METHODS

### Patients

The Hopkins Lupus Cohort is a longitudinal, prospective cohort of patients whose diagnosis of SLE is confirmed before study enrollment.^15^ Patient history, laboratory testing, and clinical information relevant to the classification of SLE and Systemic Lupus International Collaborating Clinics / American College of Rheumatology Damage Index (SLICC/ACR SDI) are recorded at the time of cohort entry.^16^ Cohort patients are followed quarterly at a minimum, and clinical information and laboratory testing are updated at subsequent clinical visits. The Hopkins Lupus Cohort is approved yearly by the Institutional Review Board of the Johns Hopkins University School of Medicine. We identified patients who either had a history of pericarditis at the time of cohort entry or were diagnosed with pericarditis during cohort participation utilizing the criterion described below.

### Diagnostic criteria for pericarditis

Pericarditis was diagnosed using the Safety of Estrogens in Lupus Erythematosus National Assessment – SLE Disease Activity Index (SELENA-SLEDAI).^17^ One of the following criteria was necessary to be present for the diagnosis of pericarditis: (1) pericardial pain, (2) auscultation of pericardial rub, (3) presence of pericardial effusion on imaging, and (4) electrocardiogram (EKG) confirmation.^17,18^ Clinical information was examined for all follow-up encounters after the first episode of pericarditis. “Recurrent” pericarditis was defined as pericarditis that occurred at least six weeks after the first recorded episode. Multiple recurrent episodes were identified if pericarditis occurred at least six weeks apart.

### Statistical analysis

The analysis was based on cohort observations that occurred after the patient’s first episode of pericarditis. For those with a history of pericarditis prior to cohort entry, all cohort follow-up encounters were included. Cohort data were reformatted into a data set with one record for each person-month of follow-up and used to calculate the rate of reoccurrence per person-month given various patient characteristics or exposures. For ease of interpretation, rates per person-month were converted to rates per person-year of follow-up. Rate ratios, confidence intervals, and p-values were calculated using pooled logistic regression (a form of discrete survival analysis) as implemented using generalized estimating equations (to account for the fact that some participants experienced more than one recurrence).^19^ Analyses were performed using SAS 9.4 (SAS Institute, Carey, North Carolina, USA).

## RESULTS

### Patient characteristics

Among the Hopkins Lupus Cohort (n=2931), 590 patients with a history of pericarditis were included in analysis. Of these 590 patients, 451 (76%) patients experienced their first episode of pericarditis prior to cohort entry, and the remaining 139 patients experienced their first episode while in the cohort. The majority of patients were women (n = 535, 91%) and identified as Black (n = 303, 51%) (Table 1). The median duration of cohort follow-up observed following the initial acute pericarditis episode was 6.7 years (IQR: 2.5 – 13.6).

**Table 1.** Demographics and Clinical Characteristics of Patients with Pericarditis in the Hopkins Lupus Cohort.

|  | **All**<br>**(N = 590)** |
| --- | --- |
| Age at first episode of pericarditis (years), no. (%) |  |
| < 30 | 257 (44) |
| 30-44 | 208 (35) |
| 45-59 | 102 (17) |
| ≥ 60 | 23 (4) |
| Female sex, no. (%) | 535 (91) |
| Race, no. (%) |  |
| White | 253 (43) |
| Black | 303 (51) |
| Other | 34 (6) |
| Onset of pericarditis, no. (%) |  |
| < 1990 | 106 (18) |
| 1990 – 1999 | 165 (28) |
| 2000 – 2009 | 215 (36) |
| ≥ 2010 | 104 (18) |
| Duration of follow-up after onset of pericarditis<br>(person-years), no. (%) |  |
| < 5 | 260 (44) |
| 5 – 10 | 115 (19) |
| 10 – 15 | 91 (15) |
| 15 – 20 | 55 (9) |
| ≥ 20 | 69 (12) |
| Recurrent episodes of pericarditis, no. (%) |  |
| 1 | 61 (10) |
| 2 – 4 | 46 (8) |
| 5 – 12 | 13 (2) |

### Overall rate of reoccurrence

We observed a total of 5277 years of follow-up during which time there were 278 recurrences of pericarditis for a rate of 0.053 per person-year of follow-up (95% CI 0.047, 0.059). These events occurred in 120 different patients (20%), and 59 (49%) of those patients experienced more than one episode of recurrence.

### Diagnostic criteria for pericarditis

Patients were diagnosed with pericarditis based on the SLEDAI criteria, which required the presence of at least one of the following: pericardial chest pain, pericardial friction rub on auscultation, evidence of effusion, or findings on EKG. Most patients were diagnosed based on symptomatic reports of pericardial chest pain (n = 567, 96%). EKG or imaging studies were performed on a small subset of patients: EKG was conducted in 1 (0.2%), while transthoracic echocardiogram (TTE) or computed tomography (CT) imaging findings demonstrated a pericardial effusion in 19 (3%) and 1 (0.2%) patients, respectively. Importantly, when diagnostic data was collected in conjunction with symptomatic reports of chest pain, the clinical diagnosis of pericarditis was confirmed 100% of the time.

### Risk factors for recurrent pericarditis

Demographic, clinical, serologic, and treatment characteristics were studied in univariate analysis to determine their association with recurrence among patients with pericarditis (Table 2). Sex did not significantly impact recurrence rates. Black race was associated with an increased risk of recurrent pericarditis (RR 1.72 (0.99, 2.97), *P* = 0.054), and older age was associated with lower rates of recurrent pericarditis (40-49 years [RR 0.58 (0.37, 0.90), *P =* 0.016] or 50-59 years [RR 0.25 (0.12, 0.53), *P* <.001] vs. 18-39 years). Recurrence was more likely to occur within the first year from onset, after which the likelihood of recurrence decreased (1-3 years vs. <1 RR 0.50 (0.32, 0.79), *P =* 0.003).

**Table 2.**
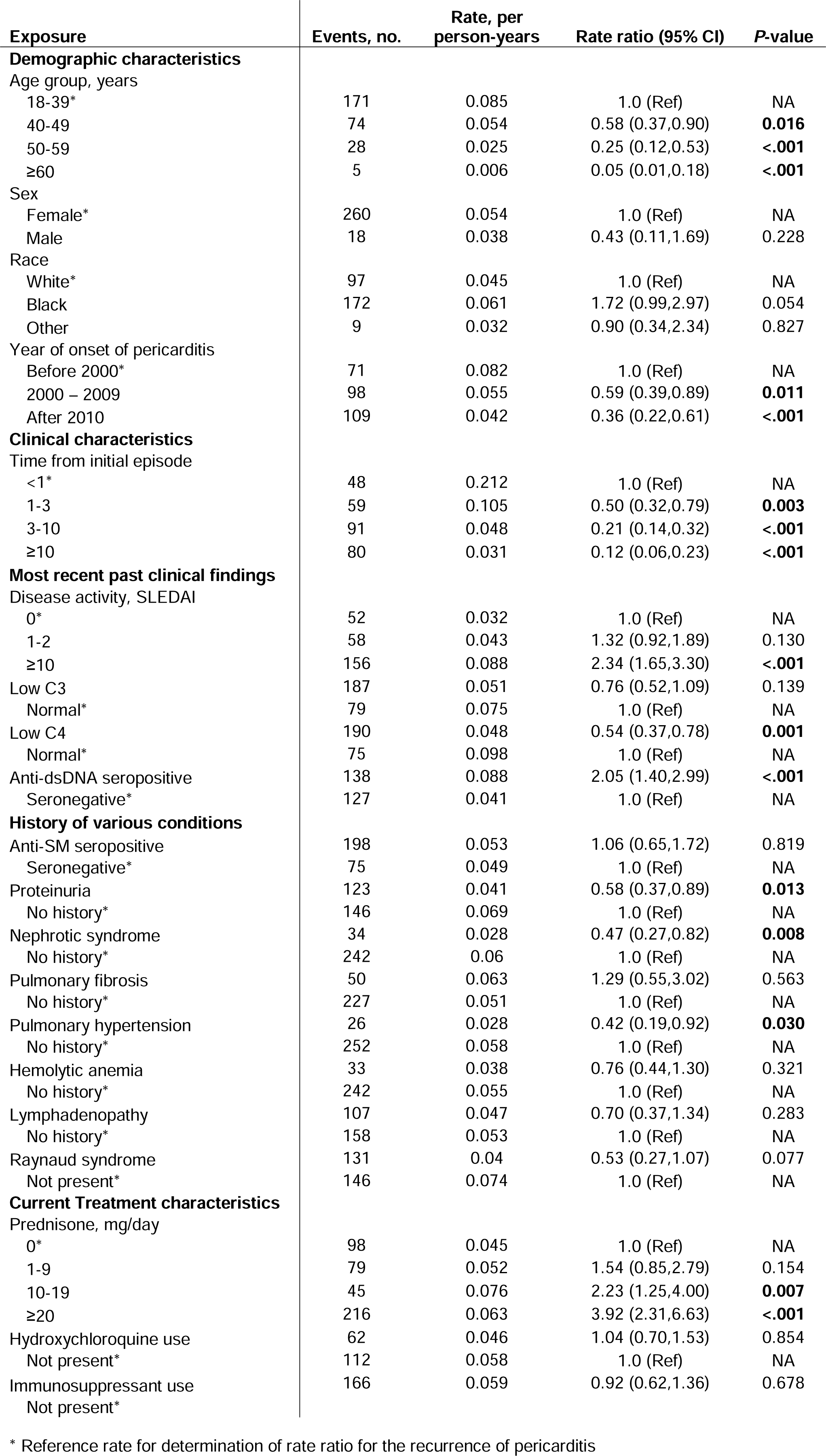
Rates of recurrent pericarditis by demographic, clinical, and treatment characteristics.

We evaluated the association of SLE disease activity with the risk of recurrence. We found that patients with active lupus (SLEDAI scale score ≥3) experienced significantly increased rates of recurrence (RR 2.34 (1.65, 3.30), *P* <.001). Systemic SLE activity beyond the pericardium was associated with a risk of recurrence. Interestingly, patients with renal involvement, such as proteinuria and nephrotic syndrome, as well as those with pulmonary hypertension, had decreased recurrence rates (Table 2). Hypocomplementemia (C3 and C4) was associated with increased rates of recurrence. However, a statistically significant association was limited to C4 (Normal vs. low C4, RR 0.54 (0.37, 0.78), *P* = 0.01) rather than reduced C3 (Normal vs. low C3, RR 0.76 (0.52, 1.09), *P* = 0.14). Seropositivity for double-stranded DNA (dsDNA) antibodies was associated with increased recurrence (RR 2.05 (1.40, 2.99), *P* <.001).

Finally, the therapies of SLE patients with recurrent pericarditis were assessed. Treatment with oral prednisone at any dose was associated with increased rates of recurrence. Furthermore, a dose-dependent association with recurrence was observed as the greatest rates of recurrence were observed with the highest daily doses of prednisone (≥20mg) (RR 3.92 (2.31, 6.63), *P* <.001). The use of hydroxychloroquine and other immunosuppressants did not affect recurrences, and rates of pericarditis were lower after the year 2000. (Table 2).

To gain further insights into our findings, we performed a multivariate Cox hazard model incorporating covariates identified as significant in the univariate analyses (*P* < 0.05) (Table 2). The model identified younger age (< 40 years), time from first episode (< 1 year), daily prednisone use (≥ 20mg), and clinically active disease (SLEDAI ≥3) as independent predictors of recurrent pericarditis (Figure 1). Other variables that were found to be associated with recurrence in the univariate analyses (low C4, anti-dsDNA, proteinuria, and history of pulmonary hypertension) were not significantly associated with recurrence after adjustment for other variables in the multivariable model.

**Figure 1.**
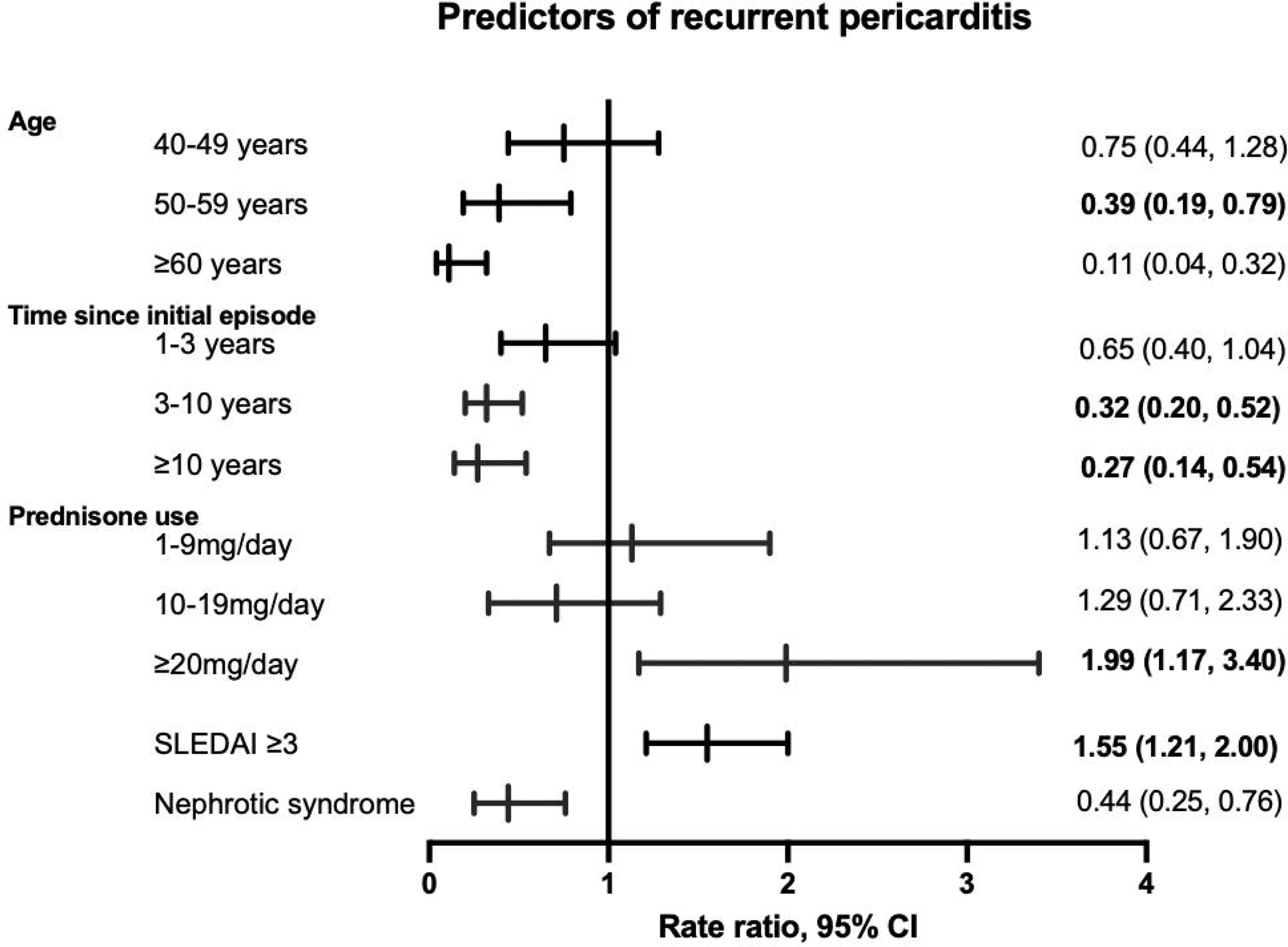
Results of multivariate analyses demonstrating predictors of recurrent pericarditis among patients with SLE. Reference rates for the determination rate ratios are summarized in Table 2.

## DISCUSSION

In this study, we present a prospective analysis of recurrent pericarditis among SLE patients. The rate of recurrent pericarditis among patients with SLE was 0.05 per year (120/590 patients, 20%). We found that whereas clinically active SLE and younger age are predictors of recurrence, systemic involvement beyond the pericardium, such as the renal system (e.g., nephrotic syndrome and proteinuria) and pulmonary hypertension, may lead to decreased recurrence. Importantly, prednisone use led to an increased risk of disease recurrence. These findings expand our understanding of lupus pericarditis, underscore the need to reconsider using oral prednisone to treat SLE flares involving the pericardium and identify the need for further investigation of treatments for lupus pericarditis.

SLE is a complex disease characterized by broad immune dysregulation that is considered the result of autoantibody production from irregularly activated and differentiated B lymphocytes and abnormal interferon activity.^20^ Similarly, the pathogenesis of recurrent pericarditis in the general population has been presumed to be immune-mediated.^8^ Given the increased potential for auto-immunity among patients with SLE, we hypothesized that patients with SLE might experience an increased rate of recurrence following an acute episode of pericarditis. Surprisingly, we found that the rate of pericarditis recurrence among patients with SLE (20%) was lower than that reported in the general population (∼33%).^4,18^

Prior studies have examined risk factors for acute pericarditis among patients with SLE. *Ryu et al.* reported predictors that include African-American ethnicity, anti-smooth muscle, anti-DNA, hemolytic anemia, proteinuria, and Raynaud’s phenomenon.^3^ However, to the best of our knowledge, there have been no studies to date that have investigated the risk factors for recurrence. We found that elevated SLEDAI scores, younger age at disease onset, recurrence occurring within one year, and higher daily doses of prednisone were independently associated with a two- to three-fold increased recurrence rate. This suggests that pericarditis recurrence reflects both patient characteristics and treatment choices. Further, these findings suggest that specific subgroups of patients with pericarditis may benefit from close monitoring and early initiation of therapies proven to be effective for recurrent pericarditis, especially as recurrence is more likely to occur within one year of the onset of pericarditis.^21^

The medical management of pericarditis for patients with SLE differs from that for the general population. Although parenteral administration of glucocorticoids may be preferred to prolonged oral administration to mitigate the risk of complications (e.g., avascular necrosis), oral glucocorticoids are still considered for the treatment of SLE flares, including pericarditis.^22,23^ In contrast, colchicine and non-steroidal anti-inflammatory drugs (NSAIDs) are preferred first-line agents for the treatment of acute pericarditis in the general population.^4,24,25^ In fact, the Investigation on Colchicine for Acute Pericarditis (ICAP) trial demonstrated that glucocorticoid use is an independent risk factor for recurrence.^25^ Furthermore, a meta-analysis of 7 studies, which included 471 patients, confirmed these findings: not only did steroids increase the risk of recurrent pericarditis, but low-dose steroids also conferred lower odds of recurrence compared to high-dose steroids (OR 0.29, 95% CI 0.13, 0.66).^26^ In the Hopkins Lupus cohort, we observed a dose-related effect of oral prednisone treatment on the risk of pericarditis recurrence similar to that in the general population, which was confirmed in the multivariate analysis. This suggests that oral prednisone should be avoided whenever possible in patients with SLE and a history of pericarditis. Further studies will be needed to understand the role of colchicine and IL-1-blocking antibodies in treating recurrent pericarditis in SLE patients.

The use of the SLEDAI criteria to diagnose pericarditis is a limitation of this study. The SLEDAI definition of pericarditis is, in fact, less stringent than that used by cardiology associations, such as the European Society of Cardiology (ESC).^18,22^ According to the SLEDAI definition, only one of the following criteria is necessary to be present for diagnosis: (1) pericardial pain, (2) auscultation of pericardial rub, (3) presence of pericardial effusion on imaging, and (4) EKG confirmation.^17,18^ Within our study cohort, patient-reported pericardial chest pain was used most often to diagnose pericarditis. Importantly, we found that all patients (n = 20) who were further evaluated via modalities, including TTE, CT chest imaging, or EKG, demonstrated findings indicative of pericarditis, thereby fulfilling both SLEDAI and ESC criteria for the diagnosis of pericarditis. This suggests that clinical criteria alone may be a robust tool for diagnosing pericarditis among patients with SLE.

To conclude, our study describes the rate of recurrent pericarditis among patients with SLE and describes risk factors associated with recurrence. Our data suggests that, despite the common practice to use prednisone to treat SLE flares, the use of oral corticosteroids should be avoided for those with a recent history of pericarditis. Future studies within this unique population will be needed to determine the most effective treatment method for pericarditis, the most common cardiac complication of systemic lupus erythematosus.

## Data Availability

All data produced in the present study are available upon reasonable request to the authors.

## Author contributions

Drs. Luigi Adamo and Andrea Fava had full access to all the data in the study and took responsibility for the integrity of the data and the accuracy of the data analysis. Study concept and design: Luigi Adamo, Andrea Fava, and Michelle Petri. Acquisition, analysis, and interpretation of data: Yoo Jin Kim, Jana Lovell, Alaa Diab, Laurence S. Magder, Daniel Goldman, Michelle Petri, Andrea Fava, and Luigi Adamo. Drafting of the manuscript: Yoo Jin Kim, Jana Lovell, Luigi Adamo, and Andrea Fava. Critical revision of the manuscript for important intellectual content: Yoo Jin Kim, Jana Lovell, Laurence S. Magder, Daniel Goldman, Michelle Petri, Andrea Fava, and Luigi Adamo. Statistical analysis: Laurence S. Magder.

## Funding/support

This study was supported in part by NHLBI grants 5K08HLO145108-03 and 1R01HL160716-01, awarded to Dr. Luigi Adamo, and NHLBI grant T32-HL007227, awarded to Dr. Jana Lovell. The Hopkins Lupus Cohort is supported by R01DK134625, awarded to Dr. Andrea Fava.

## Financial disclosures

The authors have no financial disclosures immediately relevant to this manuscript to disclose. Dr. Adamo discloses the following other relationships: consultant for Kiniksa Pharmaceutical and co-founder of i-Cordis, LLC, a start-up company focused on developing immunomodulatory small molecules to treat heart failure. All the other authors have no financial interests to report.

## Notes

### Funding Statement

This study was supported in part by NHLBI grants 5K08HLO145108-03 and 1R01HL160716-01, awarded to Luigi Adamo, and NHLBI grant T32-HL007227, awarded to Dr. Jana Lovell. The Hopkins Lupus Cohort is supported by R01DK134625, awarded to Dr. Andrea Fava.

### Author Declarations

The IRB of Johns Hopkins University gave ethical approval for this work.

